# Patient-Specific Connectomic Models Correlate With, But Do Not Predict, Outcomes in Deep Brain Stimulation for Obsessive-Compulsive Disorder

**DOI:** 10.1101/2021.04.15.21255580

**Authors:** Alik S. Widge, Fan Zhang, Aishwarya Gosai, George Papadimitrou, Peter Wilson-Braun, Magdalini Tsintou, Senthil Palanivelu, Angela M. Noecker, Cameron C. McIntyre, Lauren O’Donnell, Nicole C.R. McLaughlin, Benjamin D. Greenberg, Nikolaos Makris, Darin D. Dougherty, Yogesh Rathi

## Abstract

**Background:** Deep brain stimulation (DBS) of the ventral internal capsule/ventral striatum (VCVS) is an emerging treatment for obsessive-compulsive disorder (OCD). Recently, multiple studies using normative connectomes have correlated DBS outcomes to stimulation of specific white matter tracts. Those studies did not test whether these correlations are clinically predictive, and did not apply cross-validation approaches that are necessary for biomarker development. Further, they did not account for the possibility of systematic differences between DBS patients and the non-diagnosed controls used in normative connectomes.

**Methods:** We performed patient-specific diffusion imaging in 8 patients who underwent VCVS DBS for OCD. We delineated tracts connecting thalamus and subthalamic nucleus (STN) to prefrontal cortex via VCVS. We then calculated which tracts were likely activated by individual patients’ DBS settings. We fit multiple statistical models to predict both OCD and depression outcomes from tract activation. We further attempted to predict hypomania, a VCVS DBS complication. We assessed all models’ performance on held-out test sets.

**Results:** No model predicted OCD response, depression response, or hypomania above chance. Coefficient inspection partly supported prior reports, in that capture of tracts projecting to cingulate cortex was associated with both YBOCS and MADRS response. In contrast to prior reports, however, tracts connected to STN were not reliably correlated with response.

**Conclusions:** Patient-specific imaging and a guideline-adherent analysis were unable to identify a tractographic target with sufficient effect size to drive clinical decision-making or predict individual outcomes. These findings suggest caution in interpreting the results of normative connectome studies.

## Introduction

Deep brain stimulation (DBS) is an emerging approach to treatment-resistant mental disorders (1–3), but response rates in formal clinical trials are mixed (1,4–7). More reliable outcomes might be achieved by improving anatomic targeting. As psychiatric disorders are increasingly understood as network disorders (8,9), psychiatric DBS is moving away from using a single nucleus/structure as the target and towards attempts at affecting networks (1,10–12). There is particular enthusiasm for identifying target networks through diffusion tractography, which may enable DBS electrode placement to be customized to individual patients’ anatomy. Although there is controversy over how accurately tractography reconstructs white matter anatomy (13,14), remarkable early results have been reported from DBS placement based on that imaging (10). Further, there are multiple tools available to model the interaction of DBS electric fields and targeted tracts (15–17). These tools could replace trial-and-error DBS programming with a mathematically optimal approach to activating desired pathways while minimizing off-target effects (18). That could overcome the difficulty of correctly programming stimulation, a likely driver of inconsistent clinical outcomes (1,4,19).

To realize that promise, we need to know which tracts should/should not be stimulated. For DBS of the subcallosal white matter for depression, multiple groups have settled on a specific white matter confluence and are studying it prospectively (with varying clinical outcomes (10,20)). For obsessive-compulsive disorder (OCD), a consensus may also be emerging. A theory linking OCD to dysfunction in cortico-striato-thalamic connectivity (21,22) has led to a focus on white matter tracts linking prefrontal cortex (PFC) to striatum, basal ganglia, and thalamus. Retrospective studies from multiple institutions have implicated tracts to/from dorsolateral PFC (23,24), ventrolateral PFC (12,25,26), and anterior cingulate (12,24) as potentially important in response. Recent analyses of patients implanted at two different targets correlated OCD response with a tract linking the ventral internal capsule/striatum (VCVS) and the subthalamic nucleus (STN) with the medial PFC (12,26–28). One study further suggested that capture of tracts from orbitofrontal cortex (OFC) (23) led to non-response, although a qualitative synthesis (29) suggests that effective DBS tends to activate OFC-related fibers, and OFC-directed circuits can drive compulsive behaviors in animal models (30–32).

Although promising, these prior tractographic analyses are also limited. Many used standard atlases or connectomes derived from healthy controls, comparing these maps against electric fields from patient-specific DBS placements (12,23,27,28). Individual patients, however, show dramatic variation in their white matter topography compared to atlas standards (33). Targeting maps computed using “normative” connectomes differ from those computed from patient-specific DTI images (24). Other studies used simple isotropic field models (25), or distance between electrodes and a target tract (34) which may not accurately capture the DBS-induced electric field (16,35).

Most importantly, these analyses focused on tracts that correlate with clinical response. A variable may correlate strongly with an outcome but not be able to reliably predict that outcome, e.g. if the means are separate but the tails of two distributions overlap (36–38). Best practices in biomarker research suggest explicitly building predictive models, testing those models on held-out data, and reporting predictive performance in addition to correlation (36,37,39,40). Prediction-oriented analyses might better answer the question of whether a tractographic finding can be used as a programming target, i.e. whether it has strong predictive accuracy at the single-patient level (41).

Here, we address these limitations through an explicit attempt to predict single-patient response to DBS for OCD at the VCVS target, based on more precise field modeling approaches and using patient-specific tractography. We replicate in part prior studies’ findings that cingulate, medial PFC, and lateral PFC tracts are correlated with clinical response, but we show that these correlations do not provide strong clinical predictive power, and in some cases we identify correlations that contradict earlier reports.

## Methods

### Study Population and Clinical Treatment

Participants were 6 patients who enrolled in a clinical trial (NCT00640133) of VCVS DBS for OCD (42), plus 2 who received VCVS DBS for OCD under a Humanitarian Device Exemption. All patients received Medtronic model 3387 DBS leads, with the most ventral contact targeted to the ventral striatal grey matter. The Institutional Review Boards of Massachusetts General Hospital and Butler Hospital approved the protocols and provided ethical oversight. All participants gave informed consent, explicitly including separate consent for DBS and for neuroimaging. We report here all patients who agreed to undergo imaging. We analyzed both the Yale-Brown Obsessive-Compulsive Scale (YBOCS) and Montgomery-Asberg Depression Rating Scale (MADRS), collected at visits approximately 2-4 weeks apart by a trained rater.

### Imaging and Patient-Specific Tractography

Pre-operative MRI data were acquired on a 3T Siemens TimTrio scanner. Diffusion MRI (dMRI) scans had a spatial resolution of 2 mm (isotropic) with 10 non-diffusion weighted volumes and 60 diffusion weighted volumes, with gradient directions spread uniformly on the sphere with a b-value of 700 s/mm^2^. dMRI data were registered to pre-operative T1- and T2-weighted MRI images and post-operative CT scans using a published pipeline (43) available at https://github.com/pnlbwh/. We then performed whole brain tractography from the dMRI data, using a multi-tensor unscented Kalman filter (UKF) (44,45). The UKF fits a mixture model of two tensors to the dMRI data, providing a highly sensitive fiber tracking ability in the presence of crossing fibers (46–49). The UKF method guides each fiber’s current tracking estimate by the previous one. This recursive estimation helps stabilize model fitting, making tracking more robust to imaging artifact/noise. Another benefit of UKF is that fiber tracking orientation is controlled by a probabilistic prior about the rate of change of fiber orientation, producing more accurate tracking than the hard limits on curvature used in typical tractography algorithms. We combined the UKF with a fiber clustering algorithm to create an anatomically curated and annotated white matter atlas (48). The clustering method groups the streamlines from each patient using a spectral embedding algorithm. Each fiber cluster is matched to a tract from an *a priori* labelled atlas of the white matter derived from known connections in monkey and human brains. Fiber clustering was performed only on streamlines longer than 40 mm to annotate medium and long range tracts.

### Tract Activation Modeling

For each clinical DBS setting used in each patient, we calculated the volume of tissue activated (VTA) using a modified version of StimVision (15). Briefly, the VTAs were calculated using artificial neural network predictor functions, which were based on the response of multi-compartment cable models of axons coupled to finite element models of the DBS electric field (50). The VTAs used in this study were designed to estimate the spatial extent of activation for large diameter (5.7 µm) myelinated axons near the DBS electrode (51).

Based on theories that VCVS DBS acts by modulating circuits that run primarily in the internal capsule (14,22,29), we estimated activation of pathways linking thalamus with anterior cingulate and pericingulate cortex (ACC-PAC), dorsolateral PFC (dlPFC), ventrolateral PFC (vlPFC), dorsomedial PFC (DMPFC), medial orbitofrontal cortex (MOFC) and lateral OFC (LOFC). Pericingulate cortex includes rostral pre-cingulate cortex, but not the dorsal prefrontal cortex (such as the supplementary motor area). The atlas-guided fiber clustering algorithm (48) and a fiber clustering pipeline (52,53) guided manual delineation of fiber bundles connecting these regions to thalamus. All pathway labelings were performed by an expert neuroanatomist (Dr. Makris). Examples of the traced bundles and their intersections with DBS VTAs are shown in Figure 1A. Recent reports found that a tract connecting subthalamic nucleus (STN) to medial prefrontal cortex was strongly associated with clinical response to DBS in OCD (12,26–28). Therefore, we manually segmented the STN in each subject and extracted all fiber tracts connecting the STN with the prefrontal cortex (Figure 1B).

**Figure 1:**
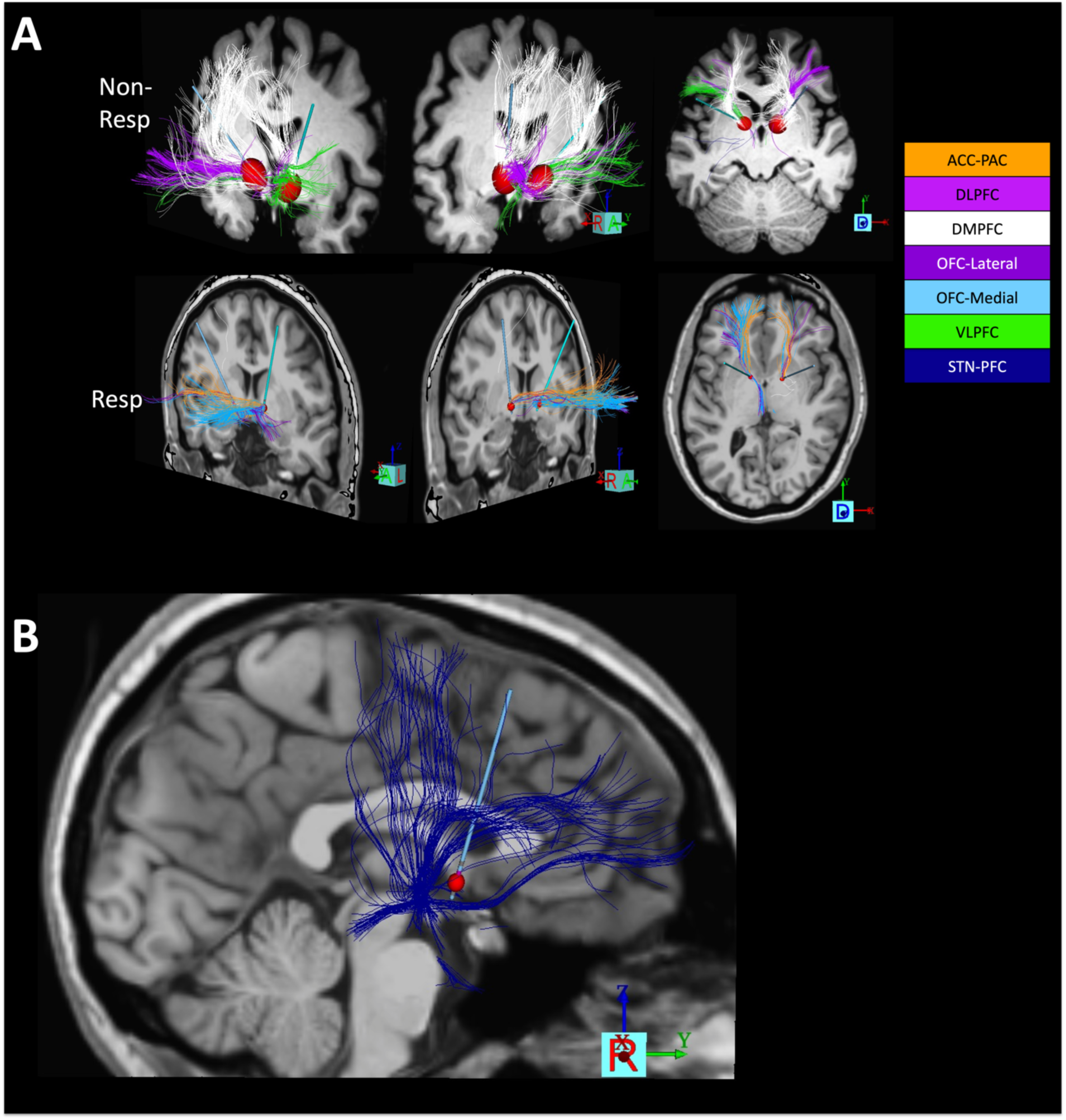
Patient-specific tractographic mapping of OCD DBS response. (A), Tract tracing and activation modeling examples. Shown are left/right oblique and axial views from one non-responder and one responder, with cortico-thalamic and cortico-STN tracts indicated by different colors. DBS leads are shown in teal and VTAs in red. In this panel, we show only tracts intersecting the VTAs for clarity. (B), Tracing of tracts between STN and frontal cortex, in the same responder as (A). To ensure capture of the tract reported in (12), we broadly traced all streamlines originating in a seed around STN and extending anterior to the central sulcus. This includes fibers coursing dorsally to motor regions, and tracts as in (12) connecting STN to ACC and medial PFC. Very few of these intersect the VTA in this patient, despite the good clinical response (YBOCS drop of 61% from baseline). To emphasize that point, this panel shows all fibers traced from the STN seed in this patient, regardless of VTA intersection.

### Data Analysis - Independent/Predictor Variables

It is unclear whether the important “dose” of DBS is activation of a sufficient number of fibers (“total fiber” model), vs. the degree to which a sub-circuit is influenced (i.e., the fraction of the overall streamlines in a tract that are within the VTA, or a “percentage” model). We calculated both and fit them as two separate models for each dependent clinical outcome (see below). We also considered the possibility that DBS response is not determined by any individual tract/pathway, but instead requires capture of multiple pathways simultaneously. We therefore added a “total activation” variable to each prediction model. For total fiber models, this variable represented the total number of streamlines activated for all tracts. For percentage models, it represented the mean percentage activation across all reconstructed tracts. We standardized all input variables to the 0-1 interval to ensure that regression coefficients were comparable between independent variables.

All models were fit and evaluated using scikit-learn (0.24.1) in Python (3.8.5). With the exception of a necessary condition analysis described below, variables were coded at the single-visit level. That is, we predicted the clinical outcome at visit T from the DBS settings programmed at visit T-1.

### Data Analysis - OCD Response

White matter pathway activation might relate tightly to the degree of clinical improvement (YBOCS as a continuous variable) or to patients’ overall well being (dichotomous responder/non-responder analysis). We thus modeled each separately. We analyzed continuous YBOCS as percentage decrease from baseline. Distribution fitting via the ‘fitdist’ package verified that YBOCS values were most compatible with a gamma distribution. We therefore predicted YBOCS improvement via an L1-regularized generalized linear regression (gamma distribution with identity link, Python package ‘pyglmnet’) and via a random forest regression with 100 trees. The dependent variable was percentage improvement in YBOCS. We compared these two approaches to assess whether conclusions might be sensitive to the model formulation. Regularized regression emphasizes selection of a small number of highly leveraged variables, which may be more helpful in defining clinical decision rules. Random forests can outperform generalized linear regression in at least some cases (54), particularly where there are nonlinearities better captured by thresholding.

We further analyzed categorical (non)response, defined as a 35% or greater YBOCS decrease from baseline (42). For these, we compared an L1-regularized logistic regression and a random forest classifier with 100 trees. A minority of visits represented clinical response (29 visits out of 165, although 5 of 8 patients were in clinical response during at least one visit). To compensate for this imbalance, we applied the Synthetic Minority Oversampling Technique (SMOTE, (55)) with 3 nearest-neighbor examples. We chose L1 regularization for both regressions because dominant models of OCD argue that dysfunction in specific cortico-striatal loops leads to symptoms (21,22) and/or that a relatively small number of fiber bundles can explain response (12,26–28). This should be reflected in clinical response being driven a small subset of tracts.

### Data Analysis - Depression Response

VCVS may have more effects on mood than on compulsivity (56), which would be reflected in better prediction of mood (MADRS) than of YBOCS. We applied the modeling pipeline used for categorical YBOCS response to categorical MADRS response, defined as a 50% or greater MADRS decrease from pre-surgical baseline. 7 out of the 165 visits met MADRS response criteria, although this again represented 5 of 8 patients.

We further assessed tractographic models’ prediction of hypomania, a known and voltage-dependent complication of VCVS DBS (57,68); details are in the Supplement.

### Data Analysis - Model Evaluation

All categorical data sets were unbalanced, and the outcome of clinical interest was always the minority class. We therefore report balanced accuracy and recall (performance for the minority class) for the categorical dependent variables. Further, we report the area under the receiver operator curve (AUC), which is suggested to be the best summary of a categorical biomarker’s performance (36,39). For continuous YBOCS prediction, we report the fraction of variance explained and the coefficient of determination (R^2^). We emphasize that R^2^ here is not the square of a correlation coefficient (36).

All metrics were calculated on a held-out test set (36,37,39,40). For each model, we held out 2 random patients from the dataset (effectively 4-fold cross-validation with resampling). This improves over leave-one-out approaches, which can overstate predictive performance (58). We left out 25% of patients, rather than visits, because data were highly autocorrelated visit-to-visit, which also falsely inflates performance (36). We then fit the predictive model on the remaining 6 patients, and we report the performance on the visit-level data from the held out patients. To prevent data leakage, the SMOTE upsampling was performed on the training set only, after the split. We obtained confidence intervals for all metrics by repeating this process over all 28 possible leave-two-out combinations, then calculating the range of performance falling within 2 standard deviations of the median performance.

We fit 16 models (4 outcomes x 2 types of model x 2 ways of expressing activation), cross-validating within each model. We interpreted the outcomes using an uncorrected 95% confidence interval to maximize power.

### Data Analysis - Predictor Importance

To detect potentially relevant tracts, we performed importance scoring on all models, regardless of whether they correctly predicted the clinical outcomes. For regression models, we computed the median and standard deviation of the regression coefficient for each tract, across all the train-test splits. For random forests, we applied permutation importance as implemented in scikit-learn. We permuted each independent variable 5 times for each of the train-test splits.

### Data Analysis - Alternative Univariate Approach

Recent papers (12,26–28) used a different approach, based on comparison of VTAs to population-scale tractography. As an additional analysis (not pre planned), we attempted a similar approach on this dataset. We calculated all linear correlations between YBOCS improvement (continuous variable) and the activation of each individual tract (either as a total fiber or percentage activation). These correlations were performed on the training set after holding out 2 random patients, consistent with (12). To test whether this approach produced more generalizable predictors of DBS response, we used the same data to fit a univariate linear regression for each independent variable, then evaluated the model performance (coefficient of determination, R^2^) on the 2 held out patients.

In a further exploratory analysis (see Supplement), we considered whether DBS outcomes depended not on the tracts activated, but the integrity of those tracts.

## Results

### Clinical Outcomes - YBOCS

The mean YBOCS improvement (considering each patient’s best time point) was 46.6%, and 5 of the 8 patients (62.5%) were clinical responders (≥35% YBOCS drop) for at least one visit.

No tract reliably predicted continuous YBOCS improvement. By all metrics, model performance was worse than chance on the held-out test set (Table 1), for both total-activation and percentage-activation models. Consistent with this, no coefficients in the regression models were above zero (i.e., the dataset mean was more reliable than any tractographic predictor). In the random forest models, the highest importance was percentage activation of fibers connecting thalamus to left OFC, but this was at chance level (change in R^2^ across models: mean 0.09, SD 0.24).

**Table 1:** Modeling outcomes for YBOCS improvement as a continuous variable. All confidence intervals include 0. Negative coefficients of determination (R^2^) imply a model that performs worse than chance.

|  | R<sup>2</sup> |  |  | Explained Variance |  |  |
| --- | --- | --- | --- | --- | --- | --- |
|  | Median | CI Lower Bound | CI Upper Bound | Median | CI Lower Bound | CI Upper Bound |
| **L1 Regression (Percentage)** | -0.194 | -1.743 | 1.355 | 0 | 0 | 0 |
| **L1 Regression (Total Fibers)** | -0.196 | -1.747 | 1.356 | 0 | 0 | 0 |
| **Random Forest (Percentage)** | -0.792 | -3.794 | 2.21 | -0.023 | -0.902 | 0.857 |
| **Random Forest (Total Fibers)** | -1.389 | -4.934 | 2.156 | -0.222 | -1.249 | 0.805 |

Similarly, no model exceeded chance for response/nonresponse prediction (Table 2). In the logistic regression, highly weighted features across models were the number (but not percentage) of activated streamlines connecting thalamus to left cingulate, lateral OFC, medial OFC, and vlPFC. Cingulate and lateral OFC streamline activation were positively associated with response, whereas medial OFC and vlPFC activation were negatively associated (Figure 2). For all of these tracts, the confidence interval for the coefficient estimated across all train-test splits included 0. These findings were sensitive to the modeling approach; the same tracts did not show median importance scores different from 0 in the random forest models. The ACC-PAC findings were corroborated by a Necessary Condition Analysis on white matter integrity (Supplementary Results).

**Table 2:** Modeling outcomes for YBOCS improvement as a categorical response. All confidence intervals include chance (0.5 for Balanced Accuracy and AUC, 0 for Recall of the minority class).

|  | Balanced Accuracy |  |  | Recall |  |  | AUC |  |  |
| --- | --- | --- | --- | --- | --- | --- | --- | --- | --- |
|  | Median | CI Lower Bound | CI Upper Bound | Median | CI Lower Bound | CI Upper Bound | Median | CI Lower Bound | CI Upper Bound |
| **L1 Logistic (Percentage)** | 0.5 | 0.5 | 0.5 | 0 | 0 | 0 | 0.5 | 0.5 | 0.5 |
| **L1 Logistic (Total Fibers)** | 0.5 | 0.093 | 0.907 | 0.571 | -0.24 | 1.383 | 0.572 | 0.281 | 0.864 |
| **Random Forest (Percentage)** | 0.46 | 0.192 | 0.728 | 0 | -0.582 | 0.582 | 0.58 | 0.368 | 0.793 |
| **Random Forest (Total Fibers)** | 0.421 | 0.193 | 0.65 | 0 | -0.527 | 0.527 | 0.588 | 0.374 | 0.802 |

**Figure 2:**
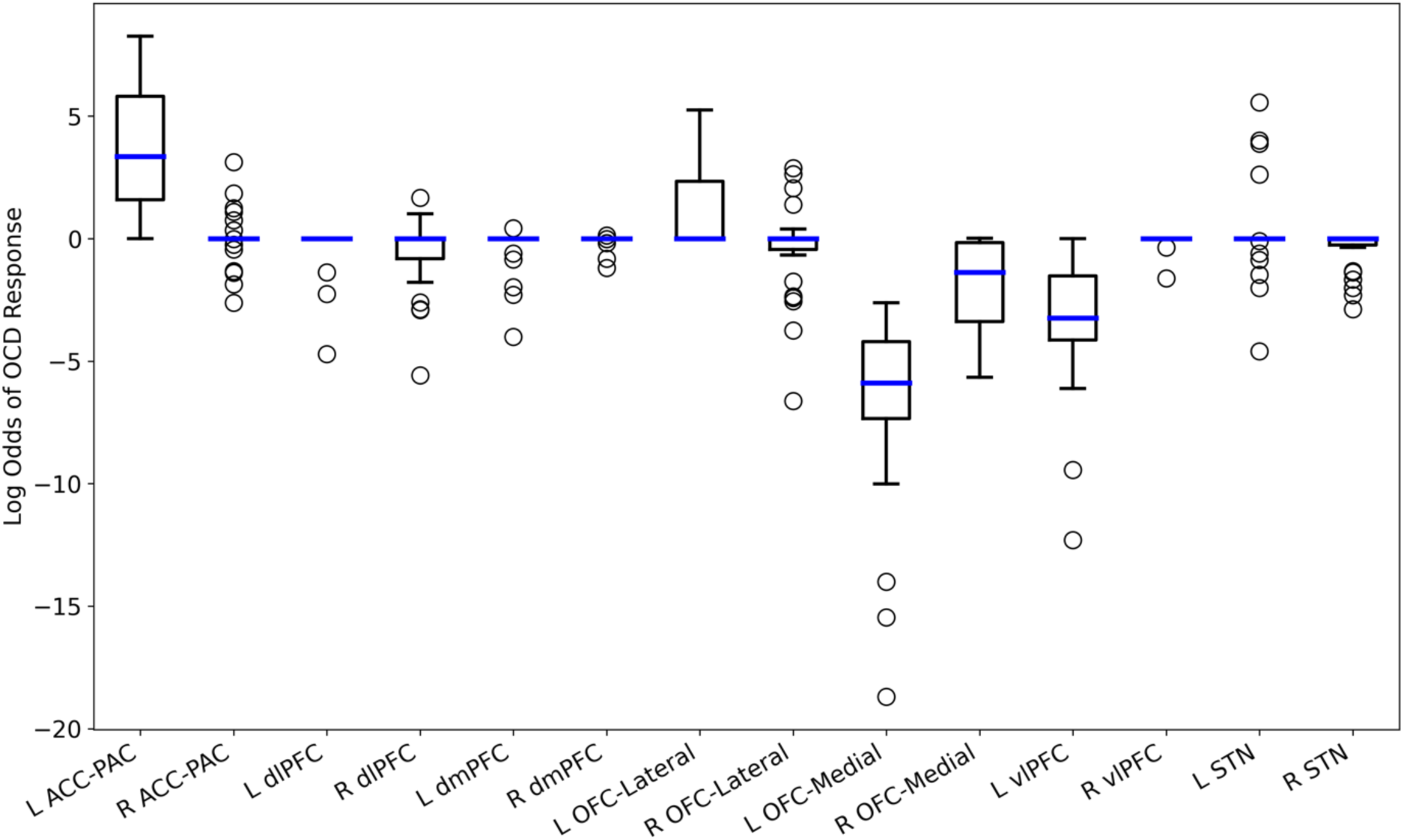
Non-zero regression coefficients across exhaustive leave-two-out cross-validation of regularized logistic regression to predict YBOCS response. All confidence intervals include 0, with left medial OFC (non-response) and left ACC (response) coming closest to significance. All reported results are for total fiber capture; percentage capture did not have non-zero coefficients in this analysis. Data are coded such that positive regression coefficients represent clinical improvement.

The alternate mass-univariate approach also did not reliably predict response on the held-out test sets (Table 3). It was concordant with the categorical response analysis in that it identified streamlines connecting the left cingulate to thalamus as correlated with response, and similarly streamlines from bilateral vlPFC as correlated with non-response. There was more discordance than similarity, however. The medial OFC tracts identified by regression were not selected in the mass univariate approach, and conversely, the mass univariate approach predicted nonresponse if tracts projecting to dlPFC were within the VTA. Further, the mass univariate approach emphasized percentage capture, while the logistic regression emphasized total fibers within a VTA. We note that tracts from STN to PFC were negatively correlated with clinical outcomes, whereas prior reports identify them as positively correlated (12,27,28).

**Table 3:** Correlations between individual fiber tracts and YBOCS response, in the style of (12), filtered to tracts whose confidence interval excludes 0 on the training sets. No such tract has clinical predictive power on held-out test sets (all R^2^ values less than 0).

| Tract | Training Set R |  |  | Test Set R<sup>2</sup> |  |  |
| --- | --- | --- | --- | --- | --- | --- |
|  | Median | CI Lower Bound | CI Upper Bound | Median | CI Lower Bound | CI Upper Bound |
| L dIPFC (Percentage) | -0.405 | -0.607 | -0.203 | -0.139 | -1.842 | 1.564 |
| L vIPFC (Total Fibers) | -0.389 | -0.591 | -0.187 | -0.268 | -2.813 | 2.277 |
| R vIPFC (Percentage) | -0.384 | -0.504 | -0.263 | -0.162 | -1.622 | 1.297 |
| L STN (Percentage) | -0.373 | -0.62 | -0.125 | -0.301 | -3.189 | 2.587 |
| L vIPFC (Percentage) | -0.366 | -0.603 | -0.129 | -0.411 | -27.119 | 26.297 |
| R dmPFC (Percentage) | -0.359 | -0.481 | -0.237 | -0.275 | -2.182 | 1.633 |
| R dIPFC (Percentage) | -0.356 | -0.542 | -0.17 | -0.14 | -1.706 | 1.426 |
| All Regions (Percentage) | -0.347 | -0.555 | -0.138 | -0.35 | -1.598 | 0.897 |
| L dIPFC (Total Fibers) | -0.332 | -0.594 | -0.07 | -0.275 | -2.149 | 1.6 |
| R STN (Percentage) | -0.321 | -0.525 | -0.118 | -0.315 | -2.149 | 1.519 |
| L dmPFC (Percentage) | -0.315 | -0.567 | -0.063 | -0.434 | -22.196 | 21.329 |
| All Regions (Total Fibers) | -0.311 | -0.553 | -0.069 | -0.421 | -1.868 | 1.026 |
| R dmPFC (Total Fibers) | -0.266 | -0.471 | -0.062 | -0.35 | -1.832 | 1.132 |
| R vIPFC (Total Fibers) | -0.266 | -0.461 | -0.071 | -0.182 | -2.991 | 2.626 |
| L ACC-PAC (Total Fibers) | 0.268 | 0.067 | 0.469 | -0.33 | -1.959 | 1.298 |
| R OFC-Lateral (Percentage) | 0.319 | 0.07 | 0.567 | -0.315 | -1.771 | 1.141 |

### Clinical Outcomes - MADRS

The mean MADRS improvement (considering each patient’s best time point) was 55.69%, and 5 of the 8 (62.5%) were responders (≥50% MADRS drop) at some point. Mood and OCD response were not linked (r=0.13 for correlation between response status on YBOCS and MADRS). Consistent with other reports (56), there were more observations of MADRS response without YBOCS than of YBOCS response without MADRS (22 vs. 4).

No model reliably predicted MADRS response above chance (Table 4). For comparison with the YBOCS analysis, we further examined the non-zero coefficients of the total-fiber regression. Capture of streamlines between right cingulate and thalamus was correlated with MADRS response, and the confidence interval for this coefficient excluded zero (Figure 3). This was not true of any other tract. Left vlPFC was associated with non-response (as it was in the categorical YBOCS analysis), but the distribution of coefficients across analyses included zero. Random forest importance scores were centered around zero.

**Table 4:** Leave-two-out prediction outcomes for categorical depression response (MADRS). No model exceeded chance accuracy on the test set (all confidence intervals include a balanced accuracy/AUC of 0.5 or recall of 0).

|  | Balanced Accuracy |  |  | Recall |  |  | AUC |  |  |
| --- | --- | --- | --- | --- | --- | --- | --- | --- | --- |
|  | Median | CI Lower Bound | CI Upper Bound | Median | CI Lower Bound | CI Upper Bound | Median | CI Lower Bound | CI Upper Bound |
| L1 Logistic (Percentage) | 0.5 | 0.5 | 0.5 | 0 | 0 | 0 | 0.5 | 0.5 | 0.5 |
| L1 Logistic (Total Fibers) | 0.5 | 0.093 | 0.907 | 0.571 | 0 | 1 | 0.572 | 0.281 | 0.864 |
| Random Forest (Percentage) | 0.46 | 0.192 | 0.728 | 0 | 0 | 0.582 | 0.58 | 0.368 | 0.793 |
| Random Forest (Total Fibers) | 0.421 | 0.193 | 0.65 | 0 | 0 | 0.527 | 0.588 | 0.374 | 0.802 |

**Figure 3:**
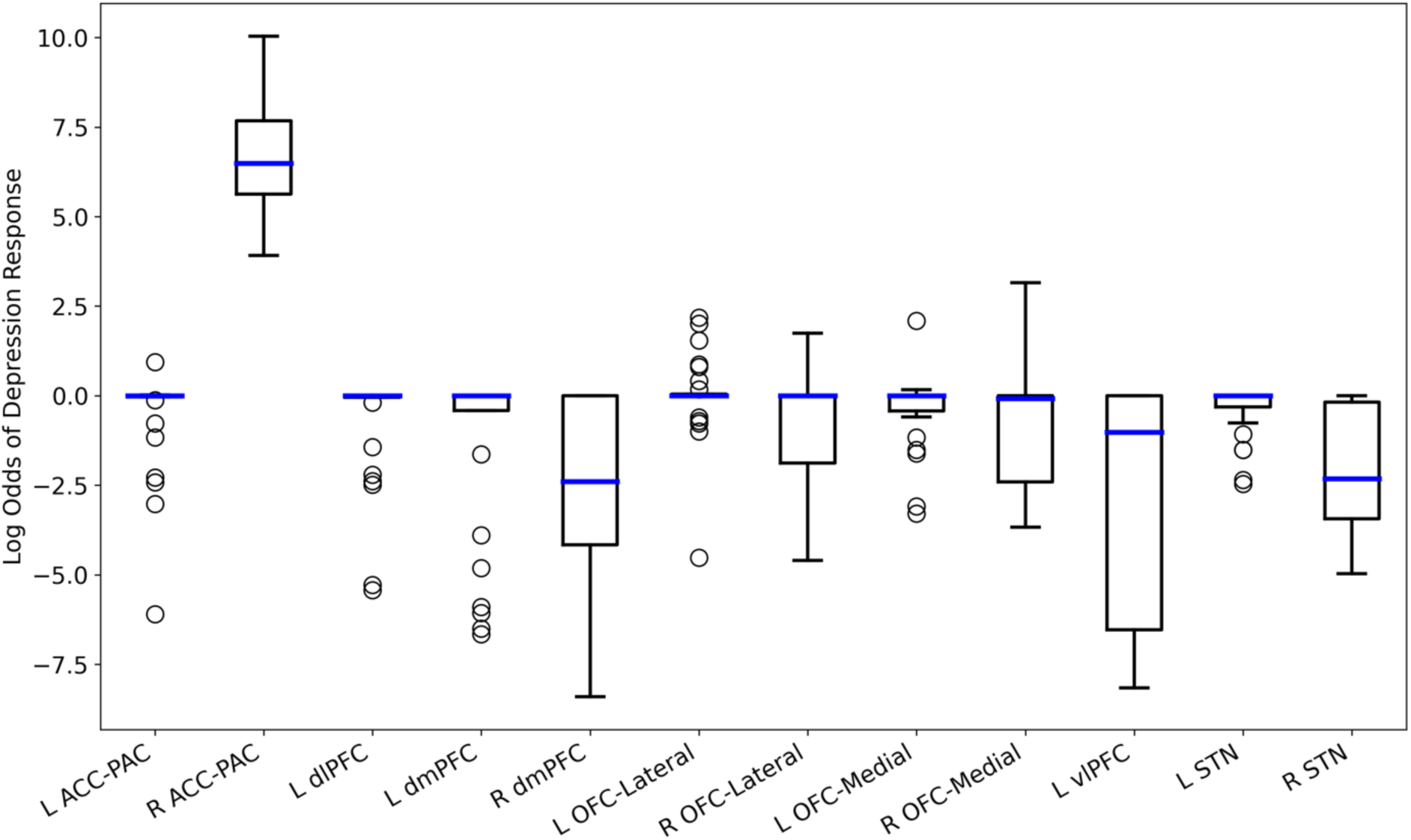
Non-zero regression coefficients across exhaustive leave-two-out cross-validation of regularized logistic regression to predict MADRS response. All confidence intervals include 0, except for the right cingulate cortex. All reported results are for total fiber capture; percentage capture did not have non-zero coefficients in this analysis.

## Discussion

Our results are both concordant and discordant with prior efforts to predict clinical OCD DBS response from tractographic modeling of cortico-striatal and cortico-basal circuits. Critically, we implemented multiple analytic steps beyond prior studies: individualized, patient-specific tracts registered to individual lead placements, activation volume calculation beyond simple electric field assumptions, consideration of multiple clinical timepoints for each patient, and formal evaluation of predictive power (as compared to measurement of correlations between activation and response or group mean differences). With this more guideline-adherent approach, we found that no tract could reliably predict clinical response or complications, whether those were considered in a continuous or categorical approach. This is likely not a surprise – we and others have highlighted that group-level significant correlations/separations often do not have clinical predictive power (36–39). In this sense, our results support calls for caution regarding the clinical role of tractography (16,41). We also showed that outcomes can be sensitive to the analytic approach – our random forest and regularized regression approaches produced very different results, even though both are commonly used approaches to prediction and variable selection.

Model inspection may offer some insight into variables for further investigation, even if pathway activation modeling approaches are not yet able to strongly predict response. Numerically, predictive power was greater (more non-zero regression coefficients after regularization) when predicting categorical rather than continuous outcomes. This may be because categorical outcomes effectively smooth out small fluctuations in continuous rating scales, fluctuations that may be primarily due to inter-rater variability or disease-unrelated variables rather than to DBS settings. The YBOCS in particular shows non-linear behavior at high scores that may exacerbate this (59). We obtained non-zero regression coefficients for models using activated fiber counts, but not for percentage-activated models, implying that it is more important to get at least a portion of a key tract within the VTA. These results also make sense in the context of our finding that the integrity (traceability) of these tracts varies greatly between patients with OCD -- a tract where response depends on tract integrity will have a large coefficient in a total-fibers model, but not in a percentage-activation model.

Our results in part support and in part diverge from a series of recent papers implicating pathways between PFC and basal ganglia as critical for OCD DBS (12,26–28). Consistent with that work, the ACC/PAC to thalamus tracts were implicated in both YBOCS and MADRS response, and were the most positively weighted in our mass-univariate approach. Our white matter integrity analysis identified the same tracts as having the largest effect size (necessity). Also similar to that prior work, we found that activation of connections to medial OFC produced numerically worse outcomes. Inconsistent with the prior work (12,26–28), we found negative correlations (in the mass univariate analysis) or null effects (in the predictive models) specifically for tracts connecting PFC to STN or vlPFC to thalamus. This again may reflect the importance of patient-specific imaging. Given that we have previously shown these tracts to have substantial inter-individual variability in their position within the internal capsule (33), and that here we note them to have similar variability in their overall integrity, a normative connectomic analysis may not reflect the actual fibers being successfully modulated in DBS cases. Alternatively, our results may highlight programming and surgical differences. These patients were implanted and programmed following the approach in (60), which emphasizes an initial search for a positive affective response. Other centers have reported very different programming algorithms (61), based more on standard anatomic positions. If response correlates with, e.g., the quality of concomitant therapy (26,62) or general clinical expertise (63), those factors will likely be strongly correlated with the programming clinician, and thus will spuriously load onto the tracts and implant locations that clinician happens to prefer. Most importantly, our results highlight the importance of applying analyses designed specifically to identify clinical predictors (36). Interestingly, we found that OFC engagement predicted worse OCD clinical response. OFC-originating components of cortico-striato-thalamic circuits are heavily emphasized in theoretical (21,22,29) and animal (30,32,64) models of OCD, and these findings may contribute to an ongoing debate over those models.

These results are tempered by three limitations. First, our sample size is small, consistent with the rarity of these patients (65). Second, imaging was not performed on a connectome-optimized scanner, and scanning at 7 Tesla (as has now become more common (66)) might identify more tracts. Third, we used relatively simple models of DBS activation. All of these add noise, reducing our ability to detect subtle correlations, particularly given DTI’s susceptibility to false positives (14). Practically, however, these limitations may not affect the clinical importance of our findings. We mitigated the lower resolution of these scans by use of an algorithm that is specifically designed to perform well in the presence of noise (45) and ensuring that our extracted tracts matched known, anatomically verified fiber bundles (48). Further, small sample sizes tend to inflate effect sizes and bias towards positive conclusions (67), not the negative result we report. Most importantly, for a tractographic result to be sufficiently reliable to inform clinical targeting/programming, it would need to have a large and clear influence on outcomes, with robustness to minor variations in analytic or clinical technique. Such a large effect would be clearly detectable and consistent across studies even at small sample sizes, like the clinical effect of VCVS DBS, which shows consistent 60-70% response rates across many small to medium cohorts (56,68–71). In that context, failure to identify a significant predictor in this small sample is relevant to both clinical practice and future study design.

Overall, our results support a growing argument that circuits linking ACC to thalamus and basal ganglia are important to VCVS DBS response. They dovetail with other work linking modulation of those circuits to increased cognitive control (72,73), a construct that is thought to be deficient in OCD (74,75). At the same time, they highlight that the current level of tractographic understanding does not have strong clinical predictive power, and that multiple confounds remain to be controlled/addressed. With multiple technologies emerging to better verify target engagement and address patient heterogeneity (1,16), that understanding will likely grow in coming years.

## Data Availability

De-identified data tables and analysis code used to produce all exhibits in this manuscript will be available at the time of journal publication at https://github.com/tne-lab.

## Data/Code Availability

De-identified data tables and analysis code used to produce all exhibits in this manuscript will be available at the time of publication at https://github.com/tne-lab.

## Acknowledgements

This work was supported by NIH grants R03MH111320 (ASW, CCM, DDD), UH3NS100548 (ASW, DDD), R01MH111917 (ASW, DDD, NMM, YR), U01U01MH076179 (BDG), P20GM130452 (NCM, BDG), K23MH100607 (NCM), and P50MH106435 (NCM, BDG). ASW further acknowledges support from the MnDRIVE Brain Conditions and Minnesota Medical Discovery Team – Addictions initiatives. All opinions and conclusions herein are those of the authors. They do not represent the views or official policy of any public or private funding body.

## Supplemental Methods

### Study Population and Clinical Treatment

In all cases, DBS implantation and programming followed the protocols described in (60). 2 patients were implanted and followed at Butler Hospital/Brown University Medical School; the other 6 were implanted and followed at Massachusetts General Hospital. All patients received Medtronic model 3387 DBS leads, with the most ventral contact targeted to the ventral striatal grey matter. All participants gave informed consent after multiple meetings with the site study teams, which explicitly included separate consent for neuroimaging.

One patient, reported in (76), declined rating visits with the study team after her first several months of treatment, due to being substantially improved and not desiring further programming. She had a series of telephone notes captured in the electronic medical record over the course of a year documenting that she was doing well with no DBS setting changes from her last programming visit. We carried her YBOCS and MADRS forward from the last available rating, despite suggestions in the clinical record that she was doing better than these scores reflect.

### Imaging Details

Pre-operative MRI data were acquired on a 3T Siemens TimTrio scanner. T1-weighted and T2-weighted images were acquired with a 1.2 mm isotropic voxel size; diffusion MRI (dMRI) scans had a spatial resolution of 2 mm (isotropic) with 10 non-diffusion weighted volumes and 60 diffusion weighted volumes, with gradient directions spread uniformly on the sphere with a b-value of 700 s/mm^2^. After DBS implantation, a postoperative CT scan was acquired at a spatial resolution of 0.43 x 0.43 x 0.63 mm^3^.

All MRI data was processed using a published pipeline (43) available at https://github.com/pnlbwh/. We applied axis alignment to the T1-weighted and T2-weighted images, and performed eddy current and motion correction for the dMRI images. T1-weighted images were skull-stripped using the Brain Extraction Toolkit (BET) (77,78). These masks were then manually checked and edited to ensure accurate brain extraction. In a similar manner, the CT scans were also masked to only retain the brain. The ANTS registration software (79) was then used to coregister the CT images to the T1-weighted images as well as the dMRI scans. Further, Freesurfer (v6.1) was run on the T1-weighted images to parcellate the brain (both cortex and subcortical structures) using the Desikan-Killiany atlas. Next, we registered the Freesurfer segmentation to the diffusion weighted images using pipeline scripts. All registrations (CT to T1-weighted, CT to dMRI, Freesurfer) were manually checked for accuracy.

During the tract tracing and annotation, fibers terminating in ACC vs. PAC could not be reliably distinguished across all subjects. We thus treated these adjacent regions as a single terminus.

We defined the STN-PFC tracts as all streamlines connecting the STN with the following Freesurfer segmented regions: caudal anterior cingulate, caudal middle frontal, lateral orbitofrontal, medial orbitofrontal, parsopercularis, parsorbitalis, rostral anterior cingulate, rostral middle frontal, superior frontal, and frontal pole. To ensure that we captured the relevant bundles, we dilated the STN by 1 voxel (∼ 2mm) in all directions (from our hand drawn segmentations) to include more tracts that might be close to the STN. These streamlines were analyzed identically to the cortico-thalamic tracts.

### Data Analysis - Hypomania

We considered the possibility that pathway modeling might be more useful for minimizing off-target effects than for determining response (23,80). The most common complication of VCVS DBS is a hypomanic-impulsive syndrome that may affect up to 50% of patients (57,68). (We refer to this as “hypomania” for conciseness, but acknowledge the substantial debate (61) over the naming and nature of this complication.) We ascertained hypomania by chart review of visit notes and case report forms. 3 patients experienced at least one hypomanic episode, over 50 total visits. We again compared L1-regularized logistic regression and random forest classification on this restricted dataset. It was not possible to perform this analysis on the full dataset, because the rarity of the complication meant that cross-validation strategies would produce training/test sets without sufficient examples. Further, it would likely be possible to achieve above-chance classification by identifying which patient contributed a given data point, which would be subtly reflected in the overall pattern of activation (i.e., there is a strong potential for data leakage).

### Data Analysis - Model Evaluation

We altered the cross-validation process for the hypomania model, because restricting the dataset to the 3 patients who experienced at least one hypomanic event made leave-2-out infeasible. We therefore cross-validated at the level of individual visits. We split the dataset with 80% of the data points as training and 20% as test set data, stratifying the split so that hypomanic events occurred in each dataset. We repeated this splitting process 1,000 times to obtain confidence intervals, and again performed all oversampling after the split.

### Data Analysis - Overall White Matter Integrity

In a further exploration, we noted that some tracts had very few streamlines in non-responders. We considered that white matter integrity might affect our results, such that patients could only respond if a given tract were sufficiently intact. For every tract in the previous analyses, we defined its integrity as the total number of streamlines traced, divided by the overall intracranial volume (to ensure that we did not simply trace more fibers in larger brains). Following the total/percentage fiber analyses above, we also created a predictor from the mean integrity across all tracked bundles. We could not correlate integrity against response at the visit level, since tract integrity does not change with DBS settings. Instead, we classified patients as overall responders if a plurality (40% or more) of their clinical visits had YBOCS scores below the 35% response threshold; 3 of the 8 met this criterion. We did not consider the first three months of the clinical course in determining this response, in order to emphasize response from DBS as opposed to lesion effect. We did not SMOTE this analysis, as the dataset was too small.

To test for a “threshold” level of integrity for DBS efficacy, we performed a Necessary Condition Analysis (NCA, (81)), as implemented in R package “NCA” (82), using the free disposal hull (CE-FDH) ceiling calculation. We separately tested the necessity of each tract and of overall (mean) integrity. We assessed significance of the resulting NCA effect sizes by 1000-fold permutation (shuffling responder/non-responder labels), with Benjamini-Hochberg false discovery rate correction.

## Supplemental Results

### Hypomania

3 patients (37.5%) experienced hypomania (6 total episodes) in this dataset. This is somewhat below the rate we reported from a different VCVS dataset (57), but consistent with the largest published cohort of this target in OCD (68).

No model was able to predict hypomania with better than chance performance (Table S5). Further, all fitted models had 0 median recall, i.e. predictive power was generally achieved by always predicting the majority (non-hypomanic) outcome. The percentage-capture regression model assigned all coefficients to 0, whereas the total-fiber model had some nonzero coefficients (Figure S4). Specifically, the left-sided PFC-STN connection was protective against hypomania. The left DLFPC-thalamus fibers had a similar but non-significant effect. Activation of right lateral OFC and STN fibers, on the other hand, predisposed towards hypomania, consistent with a prior analysis that found an association between right-sided monopolar stimulation and hypomania (57). The random forest model had no importance scores that systematically differed from zero.

**Table S5:**
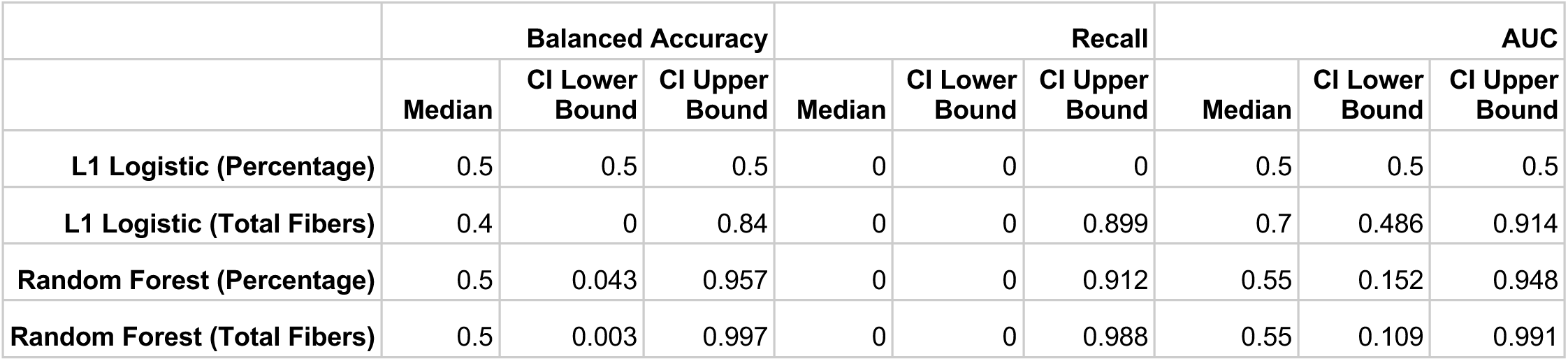
Prediction outcomes for hypomania, on a test set composed of 20% of visits from the 3 patients who had hypomanic episodes. No model consistently achieves recall > 0 or better than chance accuracy/AUC.

**Figure S4:**
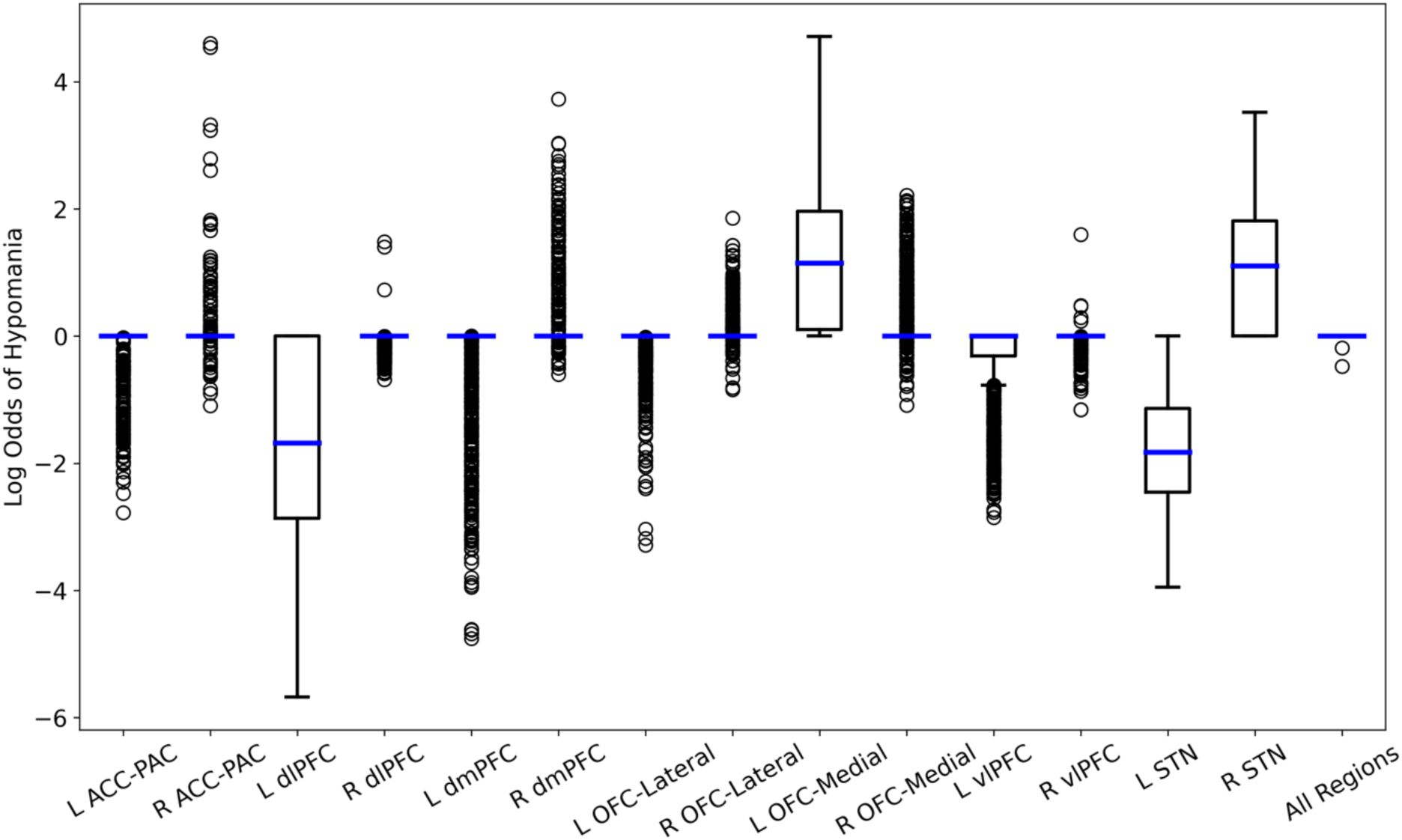
Non-zero regression coefficients for regularized logistic regression to predict hypomania. The only tract whose confidence interval excludes 0 connected left PFC to STN, and was protective against hypomania. All reported results are for total fiber capture; percentage capture did not have non-zero coefficients in this analysis. Here, positive coefficients signify greater risk of hypomania.

### White Matter Integrity

There was substantial variability in white matter integrity among individual patients and tracts (Table S2). Overall (mean) white matter integrity had a moderate effect size in the NCA (0.62). Consistent with the regression analyses, the left and right cingulo-thalamic tracts had the highest effect size of any individual tract. None of these effects reached significance even at the uncorrected level (Table S3).

**Table S6:** White matter integrity. Each cell represents the number of traced streamlines within a given tract, divided by each subject’s intracranial volume. Responder/nonresponder status is determined by the fraction of clinical visits where the YBOCS had improved 35% or more from baseline.

| Subj | L ACC-PAC | L dIPFC | L dmPFC | L OFC-Lateral | L OFC-Medial | L vIPFC | L STN | R ACC-PAC | R dIPFC | R dmPFC | R OFC-Lateral | R OFC-Medial | R vIPFC | R STN | Mean Integrity | Responder |
| --- | --- | --- | --- | --- | --- | --- | --- | --- | --- | --- | --- | --- | --- | --- | --- | --- |
| A | 0.064 | 0.280 | 0.306 | 0.017 | 0.029 | 0.414 | 0.220 | 0.155 | 0.331 | 1.083 | 0.016 | 0.010 | 0.691 | 0.115 | 0.278 | N |
| B | 0.187 | 0.506 | 1.076 | 0.012 | 0.055 | 0.279 | 0.025 | 0.160 | 0.319 | 1.718 | 0.013 | 0.033 | 0.713 | 0.025 | 0.392 | Y |
| C | 0.003 | 0.063 | 0.117 | 0.000 | 0.002 | 0.075 | 0.005 | 0.002 | 0.167 | 0.239 | 0.000 | 0.000 | 0.041 | 0.005 | 0.055 | N |
| D | 0.280 | 0.364 | 0.669 | 0.081 | 0.080 | 0.473 | 0.093 | 0.210 | 0.342 | 0.588 | 0.057 | 0.023 | 0.169 | 0.102 | 0.264 | Y |
| E | 0.148 | 0.470 | 0.834 | 0.043 | 0.089 | 0.057 | 0.105 | 0.296 | 1.079 | 0.993 | 0.008 | 0.011 | 0.418 | 0.104 | 0.350 | N |
| F | 0.194 | 0.398 | 0.871 | 0.038 | 0.092 | 0.226 | 0.054 | 0.263 | 0.302 | 1.206 | 0.051 | 0.126 | 0.688 | 0.074 | 0.347 | N |
| G | 0.311 | 0.655 | 1.327 | 0.018 | 0.042 | 0.145 | 0.567 | 0.156 | 0.413 | 0.589 | 0.016 | 0.056 | 0.149 | 0.166 | 0.342 | Y |
| H | 0.098 | 0.309 | 0.806 | 0.100 | 0.308 | 0.084 | 0.103 | 0.128 | 0.257 | 1.283 | 0.043 | 0.116 | 0.198 | 0.176 | 0.295 | N |

**Table S3:** Necessary Condition Analysis for white matter integrity predicting YBOCS response, with p-values from 1000-fold bootstrap resampling, both raw and False Discovery Rate corrected. No variable reaches corrected or uncorrected significance.

| Name | Effect Size | p | p(FDR) |
| --- | --- | --- | --- |
| Mean Integrity | 0.620 | 0.635 | 0.635 |
| L ACC-PAC | 0.595 | 0.077 | 0.475 |
| R ACC-PAC | 0.526 | 0.190 | 0.475 |
| L dIPFC | 0.509 | 0.190 | 0.475 |
| L dmPFC | 0.456 | 0.373 | 0.622 |
| R dmPFC | 0.236 | 0.635 | 0.635 |
| R OFC-Lateral | 0.225 | 0.335 | 0.622 |
| L vIPFC | 0.211 | 0.170 | 0.475 |
| R OFC-Medial | 0.182 | 0.178 | 0.475 |
| R dIPFC | 0.167 | 0.187 | 0.475 |
| R vIPFC | 0.160 | 0.635 | 0.635 |
| L OFC-Medial | 0.130 | 0.373 | 0.622 |
| L OFC-Lateral | 0.123 | 0.635 | 0.635 |
| R STN | 0.113 | 0.635 | 0.635 |
| L STN | 0.036 | 0.635 | 0.635 |

